# Age and type of task-based impact of mental fatigue on balance: Systematic review and meta-analysis

**DOI:** 10.1101/2023.11.28.23299162

**Authors:** Abubakar Tijjani Salihu, Keith D. Hill, Shapour Jaberzadeh

**Author notes:** **Corresponding Author:** Abubakar Tijjani Salihu Monash Neuromodulation Research Unit, Department of Physiotherapy, School of Primary and Allied Health Care, Faculty of Medicine, Nursing and Health Science, Monash University, Melbourne, Australia.

## Abstract

The role of cognition in balance control suggests that mental fatigue may negatively affect balance. However, cognitive involvement in balance control varies with the type or difficulty of the balance task and age. Steady-state balance tasks, such as quiet standing, are well learned tasks executed automatically through reflex activities controlled by the brainstem and spinal cord. In contrast, novel, and challenging balance tasks, such as proactively controlling balance while walking over rugged terrain or reacting to unexpected external perturbations, may require cognitive processing. Furthermore, individuals with pre-existing balance impairments due to aging or pathology may rely on cognitive processes to control balance in most circumstances. This systematic review and meta-analysis investigated the effect of mental fatigue on different types of balance control tasks in young and older adults. A literature search was conducted in seven electronic databases and 12 studies met eligibility criteria. The results indicated that mental fatigue had a negative impact on both proactive (under increased cognitive load) and reactive balance in young adults. In older adults, mental fatigue affected steady-state and proactive balance. Therefore, mentally fatigued older individuals may be at increased risk of a loss of balance during steady-state balance task compared to their younger counterparts.

## Introduction

Balance is achieved through a functioning postural control system that keeps the vertical projection of the body’s center of mass within the base of support, maintaining the body in a state of equilibrium (Boisgontier et al., 2017). Impaired postural control could result in a loss of balance potentially leading to fall (Ambrose et al., 2013; Rubenstein, 2006; Sieńko-Awierianów et al., 2018). Falls are a significant cause of mortality and morbidity across the lifespan but are more common in older people (Cho et al., 2021; Heijnen & Rietdyk, 2016; James et al., 2020; Lohman et al., 2019; Tang et al., 2021; Wang et al., 2021). Globally, fall-related, age-standardized mortality rate and years lived with disability (YLDs) were found to be 9.2 and 243 per 100,000 respectively in 2017 (James et al., 2020). In Australia, 43% of injury-related hospitalizations and 39% of injury-related deaths were due to falls between 2018-19 (Health & Welfare, 2021). In terms of healthcare costs, up to 41% (AUD 3.7 billion) of all injury-related expenditure was attributed to falls in Australia between 2015-2016 (Health & Welfare, 2020). Multiple factors play a role in falls, but most falls occur during dynamic balance activities (especially due to slip or trip during walking), implying the significance of postural balance in the etiology of falls and fall-related injuries (Ashari et al., 2021; Heijnen & Rietdyk, 2016; Wang et al., 2021).

The postural adjustments necessary for maintaining balance can be achieved through automatic or controlled (sensory motor-cognitive) processing (Boisgontier et al., 2013; Salihu et al., 2022). Typically, balance abilities that have been well learned, such as maintaining stability during quiet standing or steady-state stepping movements while walking, occur automatically through reflex activities controlled by the brain stem and spinal cord (Takakusaki, 2017). However, when encountering novel or challenging balance tasks, or in individuals with pre-existing balance impairment due to aging or pathology, the maintenance of balance is more likely to depend on cognitive processes (Bensoussan et al., 2007; Bernard-Demanze et al., 2009; Boisgontier et al., 2013; Lajoie et al., 1993; Salihu et al., 2022; Stins & Beek, 2012; Takakusaki, 2017). For instance, standing inside a moving train or walking on a slippery or rugged terrain would require cognitive process of balance control to maintain stability and prevent falls (Barra et al., 2006; Takakusaki, 2017). Likewise, individuals who are older or have sensory-motor impairments caused by specific diseases may need to rely on cognitive processes to control their balance and prevent falls, even during basic balance tasks like maintaining a quiet standing (Bensoussan et al., 2007; Bernard-Demanze et al., 2009). Considering the vital role cognitive processes play in balance control across various situations, factors that lead to temporary or permanent impairments in cognitive abilities can significantly affect balance, ultimately raising the risk of falling.

An important factor that can temporarily impair cognitive abilities potentially affecting balance and risk of falling is mental fatigue due to prolonged performance of cognitive tasks. Mental fatigue was described as a state of mental tiredness or exhaustion resulting from prolonged periods of intense or monotonous cognitive activity requiring sustained attention (Holtzer et al., 2010; Pageaux & Lepers, 2018). In addition to the subjective feeling of mental tiredness, mental fatigue is also characterized by a decrease in cognitive performance (Chen et al., 2020; Holtzer et al., 2010). Due to its effect on cognitive performance, mental fatigue may affect the cognitive processes of balance control leading to decrease stability and fall risk (Grobe et al., 2017).

However, since the cognitive involvement in balance control depend on factors such as the type or complexity of the balance task, as well as the age of the person performing the task, the effect of mental fatigue on balance may equally depend on these factors (Bernard-Demanze et al., 2009; Lajoie et al., 1993). According to Shumway-Cook and Woollacott, the functional tasks involved in day-to-day activities requires three different types of balance control namely, the steady-state, reactive, and proactive or anticipatory balance control (Shumway-Cook & Woollacott, 2007). Steady-state balance is described as the ability to control the centre of mass (COM) relative to the base of support in a predictable and nonchanging conditions (Shumway-Cook & Woollacott, 2007). Examples of tasks that require steady-state balance control include quiet standing and walking at constant velocity. On the other hand, reactive balance control involves the ability to regain stable position following unexpected perturbation (Shumway-Cook & Woollacott, 2007). For example, the restoration of stability to prevent falls following trip or slip requires proper reactive balance control ability. In the case of proactive (anticipatory) balance control, appropriate postural muscles in the leg and trunk are activated to maintain stability in advance of a potentially destabilizing voluntary movement (Shumway-Cook & Woollacott, 2007). Raising the legs one after the other to step on to a moving escalator or walking on a slippery surface, for instance, would require anticipatory muscle activity to maintain stability. Any delayed or absent muscle activity in this scenario can lead to a loss of balance.

In terms of cognitive involvement and hence the susceptibility to the effect of mental fatigue, steady-state balance control tasks, such as quiet standing, can proceed automatically with little or no cognitive involvement, making them insusceptible to mental fatigue (Brahms et al., 2022; Jacobs & Horak, 2007; Takakusaki, 2017). However, more challenging types of balance tasks requiring anticipatory and/or reactive control of balance, such as walking on a slippery surface or regaining balance following unexpected slip or trip, requires cognitive process of postural control and may therefore be affected by mental fatigue (Brahms et al., 2022; Takakusaki, 2017).

Additionally, because older adults have reduced sensitivity in the visual, vestibular, and proprioceptive systems, they are reliant on cognitive processes to control and maintain stability while performing even simple balance tasks, such as quiet standing (Boisgontier et al., 2013). Therefore, simple types of balance control tasks, such as quiet standing, which may not typically be affected by mental fatigue in young adults, can be compromised in older adults when they experience mental fatigue. Hence, it is equally important to consider age in addition to the type of balance task when investigating the effect of mental fatigue on balance.

Recently, studies investigating the effect of mental fatigue induced by prolonged cognitive performance on different types of balance tasks were conducted in different populations. However, no study summarized the findings of these studies based on the type of balance task and age of the participants in a meta-analysis. The meta-analysis by Brahms and colleagues pooled studies regardless of the type of balance task used (Brahms et al., 2022). Nonetheless, in a recent systematic review and meta-analysis, we discovered that no substantial correlation exists between performance in the different types of balance control tasks throughout the lifespan (Kiss et al., 2018). This finding suggests that balance performance is specific to individual tasks rather than being a generalized ability (Kiss et al., 2018). Therefore, pooling data from different studies without considering the type of balance task involved may not give a meaningful description of the effect of mental fatigue on balance, which reflects the task-specific nature of balance control. Moreover, the sub-group analysis based on age in the study by Brahms and associates does not answer the question of whether some types of balance tasks are affected by mental fatigue only in a particular age group, as that was not considered in the analysis (Brahms et al., 2022). Overall, the study’s failure to consider important factors in the analysis suggests that it may not have provided a comprehensive understanding of the impact of mental fatigue on balance. Consequently, its findings may not be sufficient to guide the selection of balance tasks in future studies exploring this phenomenon across various populations in clinical, occupational, or sports settings. Indeed, the authors acknowledged the need for future research that incorporates different types of balance tasks when examining the effects of mental fatigue on balance (Brahms et al., 2022). Therefore, the current systematic review and meta-analysis aimed to synthesize the available evidence relating to how different types of balance tasks are affected by mental fatigue in young and older adults.

## METHODS

The review was conducted in accordance with the Preferred Reporting Items for Systematic reviews and Meta-Analysis (PRISMA) guidelines (Moher et al., 2009).

### Data sources and search strategy

Seven electronic databases including Medline, EMBASE, Cochrane CENTRAL, Scopus, PubMed, CINAHL Plus and PsycINFO were searched for relevant studies published from inception until September 2022. The search terms used included mental fatigue OR cognitive fatigue AND balance OR postural balance OR posture OR postural stability OR postural sway. Both subject headings and keyword/free-text searches were appropriately applied in each database. An example of the search strategy for the Medline database has been provided (Table 1). Results of the database search were exported to Endnote X9 (Clarivate analytics, Philadelphia) where duplicates were removed. Manual searching of the reference list of the included studies was conducted for additional relevant studies.

**Table 1:** Example of search strategy for Medline database.

| Search | Query |
| --- | --- |
| **A** | **Medline via Ovid** |
| 1 | mental fatigue/ |
| 2 | mental fatigue.ab,kw,ti. |
| 3 | cognitive fatigue.ab,kw,ti. |
| 4 | 1 OR 2 OR 3 |
| 5 | balance.ab,kw,ti. |
| 6 | Postural balance/ |
| 7 | postural balance.ab,kw,ti. |
| 8 | posture/ |
| 9 | posture.ab,kw,ti. |
| 10 | postural stability.ab,kw,ti |
| 11 | Postural sway.ab,kw,ti. |
| 12 | 5 OR 6 OR 7 OR 8 OR 9 OR 10 OR 11 |
| 13 | 4 AND 12 |

### Study selection

After duplicate removal, the title and abstracts of the remaining studies were screened by the first reviewer, and those deemed irrelevant to the current review based on their title or content of the abstract were excluded. Full texts of the remaining studies were retrieved and subjected to the inclusion and exclusion criteria. Any uncertainty about eligibility assessment was resolved by discussion and consensus among all the authors.

### Eligibility criteria-inclusion and exclusion

Studies were included if they: 1) were written in English and published in peer reviewed journals; 2) Experimentally induced mental fatigue by asking participants to carry out prolonged cognitive tasks; 3) Investigated the effect of mental fatigue on balance by comparing the fatigue condition with a control condition using a randomized or non-randomized controlled or cross-over study design. No restriction was placed on the outcome measure used for assessment of balance in the studies. Therefore, studies using instruments for assessment of balance (e.g., using force plate) or clinical tests of balance such as the timed-up-and-go test were included. Dissertations, review articles, letters, commentaries, and conference abstracts were excluded. Studies exploring the relationship between subjective chronic mental fatigue assessed using questionnaires and balance were similarly excluded.

### Assessment of quality and risk of bias

The quality of the included studies was assessed using a modified version (Supplementary material) of the Downs and Black checklist (Downs & Black, 1998). Twenty items (1-7, 10-12, 14-16, 18-21, 23-24, 25 and 27) out of a total of 27 were included to measure study quality based on a similar approach used in some previously conducted systematic reviews (Alibazi et al., 2021; Siddique et al., 2022). The other items were excluded because they contain questions that are not relevant to the design of the studies included in this systematic review (Alibazi et al., 2021; Downs & Black, 1998; Siddique et al., 2022). Additionally, the risk of selection, performance, detection, attrition, reporting, and other biases in the included studies was assessed using the Cochrane collaboration’s tool for assessing the risk of bias as described in Chapter 8 of the Cochrane Handbook for Systematic Reviews of Interventions Version 5.1.2 (Behrangrad et al., 2021; Dissanayaka et al., 2017; Higgins et al., 2019). The risk of bias in each feature was judged as “Low risk”, ‘’High risk’’ or ‘’Unclear risk’’. Unclear risk of bias indicated either a lack of adequate information or uncertainty regarding the potential for bias in a particular feature (Behrangrad et al., 2021; Dissanayaka et al., 2017; Higgins et al., 2019).

### Data synthesis and analysis

The balance tasks used in the included studies were first categorized into steady-state, proactive (anticipatory), and reactive balance tasks based on the classification by Shumway-Cook and Woollacott (Shumway-Cook & Woollacott, 2007). Additionally, for each type of balance task, separate meta-analysis (or effect size calculations) was conducted for young and older adults. Mean, standard deviations (SD) and sample size were used to calculate effect sizes. In case some values (mean and SD) were not provided numerically, they were estimated from available plots using Plot digitizer, a Java-based software program that converts plotted values into numerical format commonly used in meta-analytical studies (Chung et al., 2016; Dissanayaka et al., 2017; Hall et al., 2021; Huwaldt, 2005; Kadic et al., 2016). In situations where increased scores on some outcome measures were associated with poorer performance, whilst increased scores on other outcome measures was associated with better performance, the mean values from one set of studies were multiplied by - 1 to correct for the difference in the direction of the scale (Higgins et al., 2019).

Wherever mean values were provided alongside SE values, the corresponding SD values were obtained using the formula SD = SE × √n (n = number of subjects). Due to mixed nature of the study design in the included studies (cross over and parallel group designs), data from the studies with cross over design were treated as if they were from parallel groups, as recommended in the Cochrane Handbook for Systematic Reviews of Interventions. Inverse variance random-effects meta-analyses were used to pool the data. Effect sizes were expressed as mean difference (MD) when the outcome measurements were made or could be converted on the same scale. Otherwise, the standardized difference in mean (SMD) was calculated. P-values of ≤ 0.05 indicated a significant effect. Forest plots with 95% confidence intervals (CIs) were reported, and standardized effect sizes were interpreted as small (˂0.1), medium (0.1–0.3) or large (˃ 0.3) (Cohen, 1988; Ghai et al., 2017). Heterogeneity was quantified using the I^2^ statistic, which can range from 0 to 100%, and was interpreted according to the rough guide provided in Chapter 9 of the Cochrane Handbook for Systematic Reviews of Interventions. For studies not included in the meta-analysis, effect sizes with 95% CI were calculated where possible. Review manager (Revman) version 5.4 was used for all analyses.

## Results

### Studies and participants

The search in the seven electronic databases yielded a total of 262 articles, which were reduced to 162 after duplicate removal (Fig. 1). The title and abstracts of the 162 articles were screened, and 139 were excluded in the process, leaving 24 articles for full-text screening. After full-text screening, twelve articles (14 data sets) met the review’s inclusion criteria and were included. The characteristics of the included studies and the participants are summarized in Table 2. Two studies investigated the effect of mental fatigue on steady-state balance in young adults (Deschamps et al., 2013; Fletcher & Osler, 2021). The effect of mental fatigue on proactive (Gebel et al., 2022; Tassignon et al., 2020; Verschueren et al., 2020) and reactive balance control (Lew & Qu, 2014; Morris & Christie, 2020; Qu et al., 2020) in the same population was investigated in three studies each. One study included both steady-state and proactive balance tasks in young adults (Hachard et al., 2020). In older adults, the effect of mental fatigue on steady state (Boolani et al., 2020; Fletcher & Osler, 2021) and reactive balance control tasks (Morris & Christie, 2020; Varas-Diaz et al., 2020) were investigated in two studies each. The effect of mental fatigue on proactive balance in older adults was investigated in one study (Nikooharf Salehi et al., 2022).

**Fig. 1.**
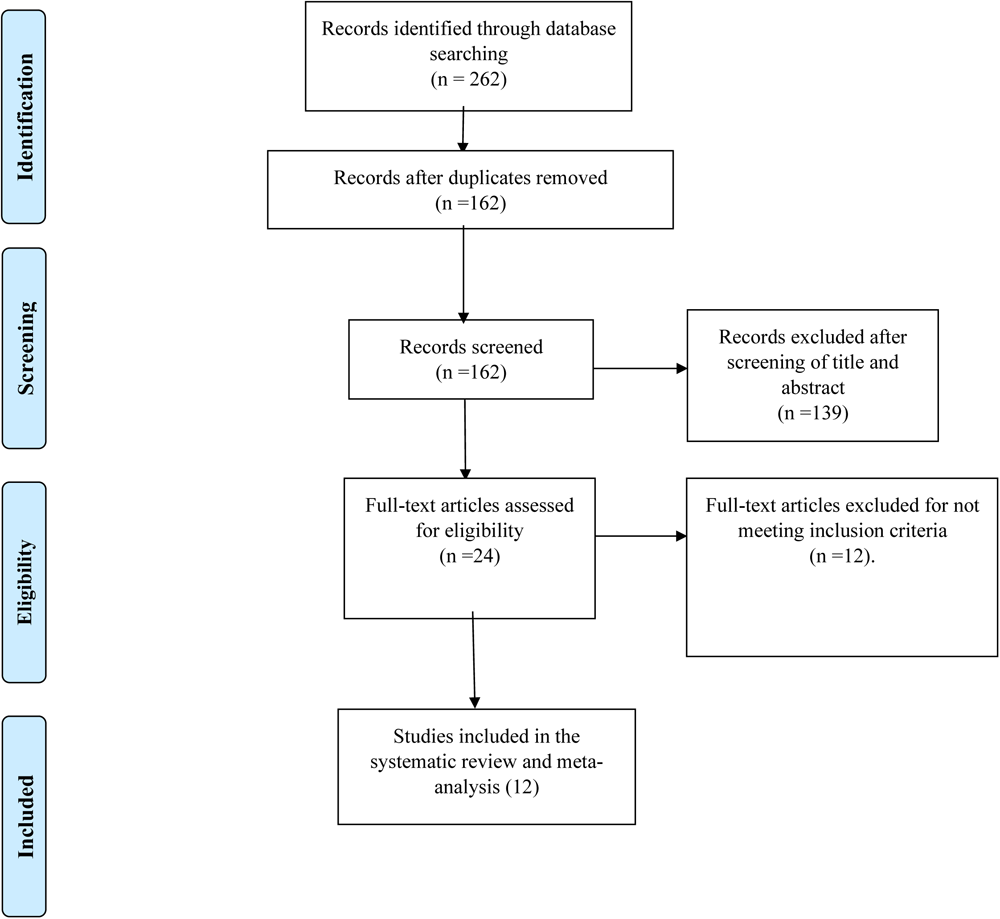
Flow diagram showing selection process of articles following PRISMA guidelines.

**Table 2:**
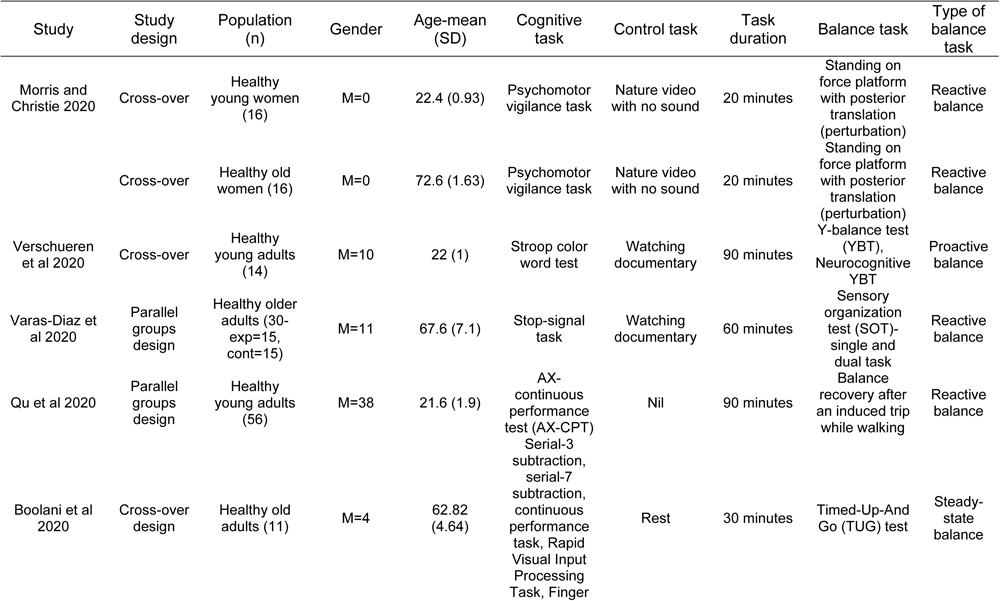
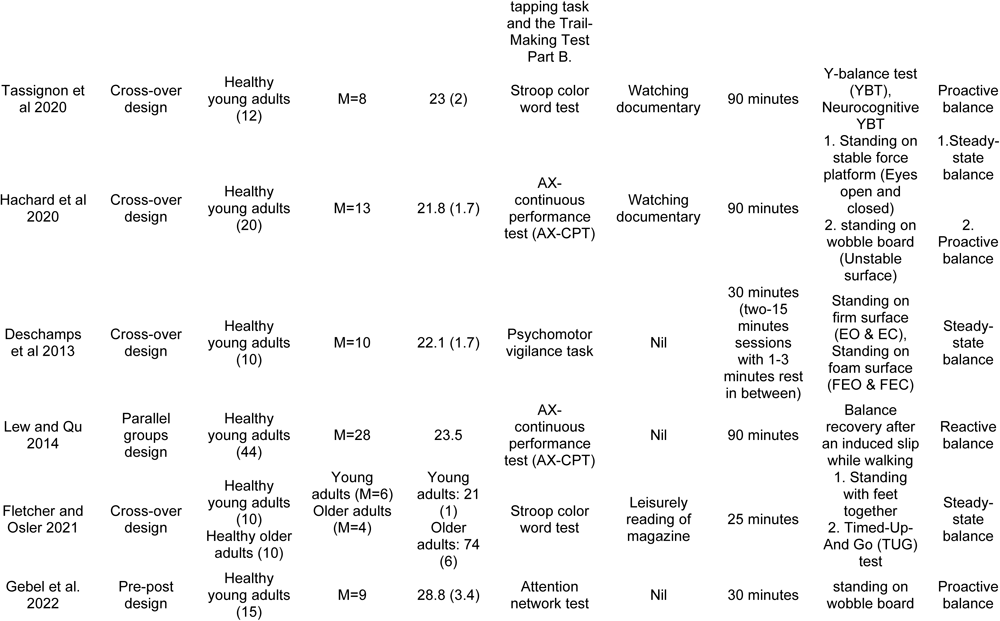
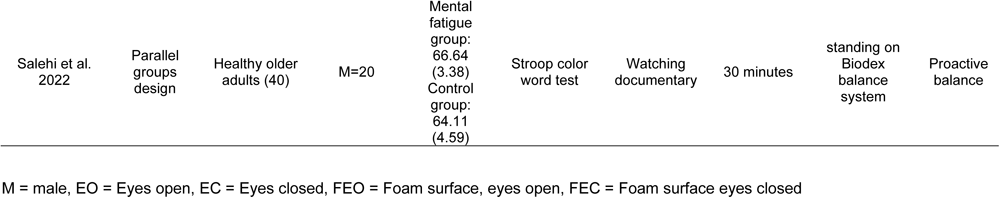
Characteristics of the included studies.

The study participants were young adults in seven studies (Deschamps et al., 2013; Gebel et al., 2022; Hachard et al., 2020; Lew & Qu, 2014; Qu et al., 2020; Tassignon et al., 2020; Verschueren et al., 2020), while two studies included both young and older adults (Fletcher & Osler, 2021; Morris & Christie, 2020). Older adults were the only participants in the remaining three studies (Boolani et al., 2020; Nikooharf Salehi et al., 2022; Varas-Diaz et al., 2020). The mean age of the young and older participants are as follows: (mean age (SD)-Young adults-22.91 (2.33) years; Older adults-67.96 (4.49) years. Mixed-gender participants were included in ten studies (Boolani et al., 2020; Fletcher & Osler, 2021; Gebel et al., 2022; Hachard et al., 2020; Lew & Qu, 2014; Nikooharf Salehi et al., 2022; Qu et al., 2020; Tassignon et al., 2020; Varas-Diaz et al., 2020; Verschueren et al., 2020). The study by Morris and Christie (2020) included only female participants, while only male participants were included by Deschamps and associates (Deschamps et al., 2013). The total number of participants in the included studies was 319 (median: n = 20, range: n = 10-56, young adults: n = 242, older adults: n = 77).

### Quality assessment and risk of bias

Out of the 12 included studies, 10 were of high quality (score of ≥70%), while the remaining two were of low quality (score of <70%) (Table 3). Regarding the risk of bias (see Figures 2 and 3), adequate random sequence generation was used in four studies, while the other three studies randomized the participants, but the method of randomization was unclear. Similarly, the allocation concealment was unclear in both the studies that used adequate random sequence and those that did not report the information about the method of randomization. Only three studies blinded the participants about the study’s aim and the fatigue hypothesis. Additionally, outcome assessment was not blinded in any of the studies, but the primary outcome was measured objectively in eight studies, and hence may not be subject to the assessor’s influence. All but one study was judged to have a low risk of attrition bias, and selective reporting was similarly noticed in only one of the included studies. Three out the six cross-over trials did not report the duration or used very short wash-out period, constituting other sources of bias in the studies. Another study included different populations of participants in the mental fatigue and control groups (people with chronic stroke versus healthy older adults), similarly constituting other sources of bias.

**Fig 2.**
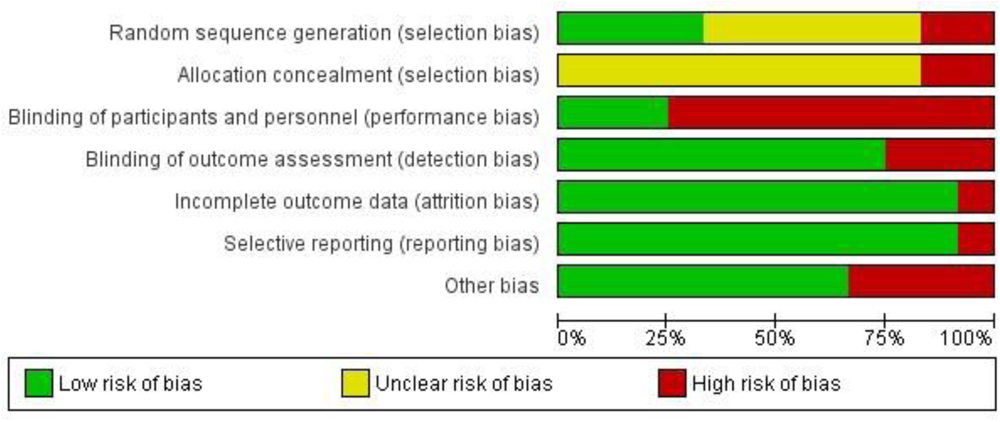
Review authors judgment about each risk of bias item presented as percentage across all included studies.

**Fig 3.**
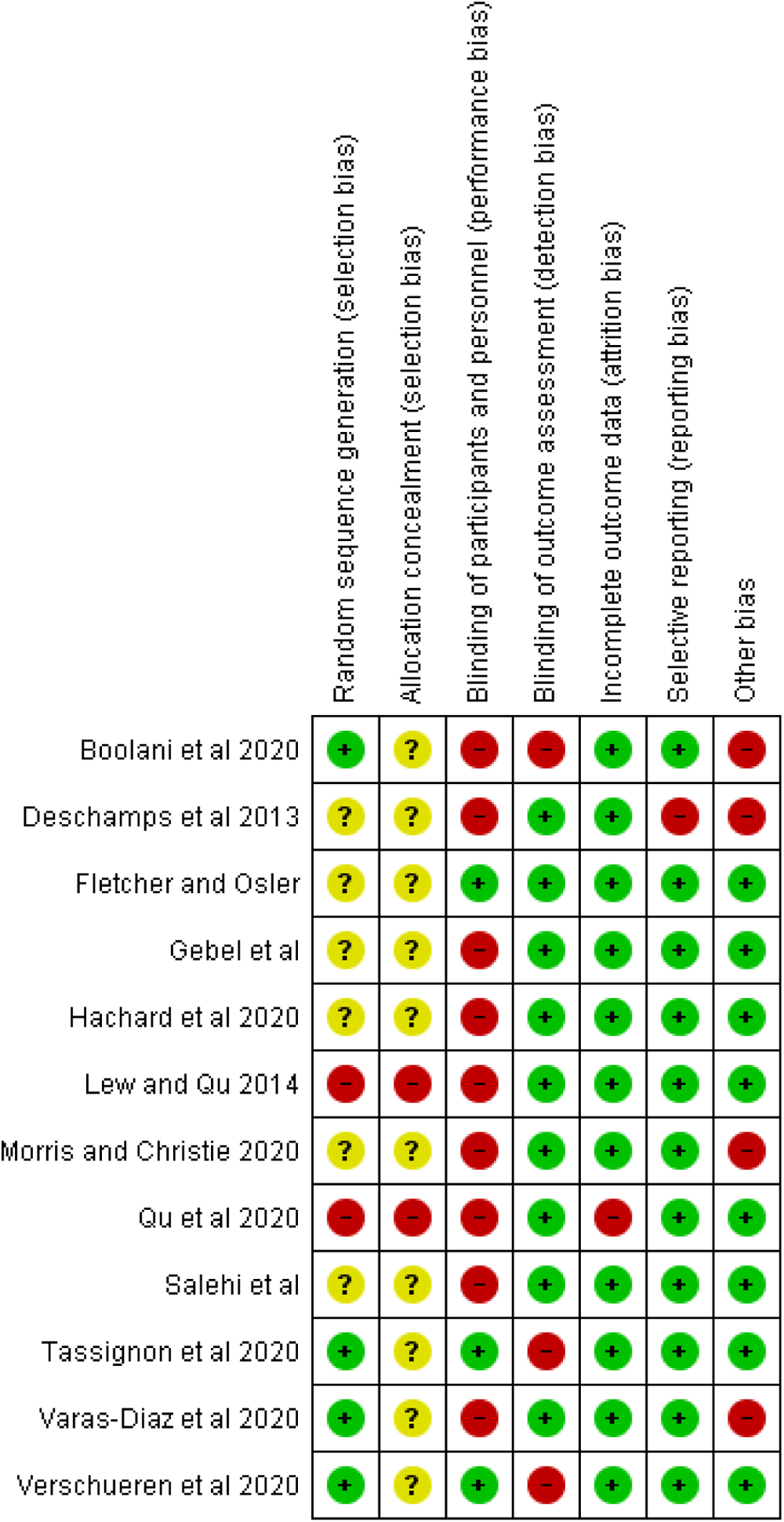
Review authors judgment about each risk of bias item for each included study.

**Table 3:** Quality assessment score for individual studies.

| Study | 1 | 2 | 3 | 4 | 5 | 6 | 7 | 10 | 11 | 12 | 14 | 15 | 16 | 18 | 20 | 21 | 23 | 24 | 25 | 27 | Total | % | Quality |
| --- | --- | --- | --- | --- | --- | --- | --- | --- | --- | --- | --- | --- | --- | --- | --- | --- | --- | --- | --- | --- | --- | --- | --- |
| Hachard et al. 2020 | 1 | 1 | 1 | 2 | 1 | 1 | 1 | 0 | 0 | 1 | 1 | 1 | 1 | 1 | 0 | 1 | 0 | 0 | 0 | 0 | 14 | 70 | HIGH |
| Fletcher and Osler 2021 | 1 | 1 | 1 | 2 | 1 | 1 | 0 | 0 | 0 | 1 | 1 | 1 | 1 | 1 | 0 | 1 | 1 | 0 | 0 | 0 | 14 | 70 | HIGH |
| Deschamps et al. 2013 | 1 | 1 | 0 | 1 | 1 | 1 | 0 | 0 | 0 | 1 | 1 | 1 | 1 | 1 | 0 | 1 | 0 | 0 | 0 | 0 | 11 | 55 | LOW |
| Gebel et al. 2022 | 1 | 1 | 1 | 1 | 1 | 1 | 1 | 0 | 0 | 1 | 1 | 1 | 1 | 1 | 0 | 1 | 0 | 0 | 0 | 0 | 13 | 65 | LOW |
| Boolani et al. 2020 | 1 | 1 | 1 | 2 | 1 | 1 | 0 | 0 | 0 | 1 | 1 | 1 | 1 | 1 | 0 | 1 | 0 | 0 | 1 | 0 | 14 | 70 | HIGH |
| Salehi et al. 2022 | 1 | 1 | 1 | 2 | 1 | 1 | 1 | 1 | 1 | 1 | 1 | 1 | 1 | 1 | 1 | 1 | 0 | 0 | 0 | 0 | 17 | 85 | HIGH |
| Morris and Christie 2020 | 1 | 1 | 1 | 2 | 1 | 1 | 1 | 0 | 0 | 1 | 1 | 1 | 1 | 1 | 0 | 1 | 0 | 0 | 0 | 0 | 14 | 70 | HIGH |
| Vaschueren et al. 2020 | 1 | 1 | 1 | 2 | 1 | 1 | 1 | 0 | 0 | 1 | 1 | 1 | 1 | 1 | 1 | 1 | 1 | 0 | 1 | 0 | 17 | 85 | HIGH |
| Tassignon et al. 2020 | 1 | 1 | 1 | 2 | 1 | 1 | 1 | 0 | 0 | 1 | 1 | 1 | 1 | 1 | 1 | 1 | 1 | 0 | 1 | 0 | 17 | 85 | HIGH |
| Qu et al. 2020 | 1 | 1 | 1 | 2 | 1 | 1 | 1 | 0 | 0 | 1 | 1 | 1 | 1 | 1 | 0 | 1 | 0 | 0 | 0 | 0 | 14 | 70 | HIGH |
| Lew and Qu 2014 | 1 | 1 | 1 | 2 | 1 | 1 | 1 | 0 | 0 | 1 | 1 | 1 | 1 | 1 | 0 | 1 | 0 | 0 | 0 | 0 | 14 | 70 | HIGH |
| Varas-Diaz et al. 2020 | 1 | 1 | 1 | 2 | 1 | 1 | 0 | 0 | 0 | 1 | 1 | 1 | 1 | 1 | 0 | 1 | 0 | 0 | 1 | 0 | 14 | 70 | HIGH |

Low-quality studies were defined as having a quality assessment score of <70%, whereas high-quality studies had a score of ≥70%

### Induction of mental fatigue

Different types of cognitive tasks and task durations were used to experimentally induce mental fatigue in the participants of the included studies. The cognitive tasks used include AX-continuous performance test (AX-CPT) (Hachard et al., 2020; Lew & Qu, 2014; Qu et al., 2020), Stroop color word test (Fletcher & Osler, 2021; Nikooharf Salehi et al., 2022; Tassignon et al., 2020; Verschueren et al., 2020) and Psychomotor vigilance tasks (Deschamps et al., 2013; Morris & Christie, 2020). One study used Stop-signal task (Varas-Diaz et al., 2020) while another one study used the attention network test (Gebel et al., 2022). The remaining one study used a combination of different cognitive tasks (Boolani et al., 2020). The task duration used ranges between 20-90 minutes (median task duration of 90 minutes/study). In five studies, the participants carried out the mentally fatiguing cognitive task for 90 minutes (Table 2). Another five studies used cognitive tasks lasting between 25 to 30 minutes, while the remaining two studies used tasks that extended for 20 and 60 minutes respectively. In most of the studies, participants watched a documentary of the same duration as the cognitive task during the control session (Hachard et al., 2020; Morris & Christie, 2020; Nikooharf Salehi et al., 2022; Tassignon et al., 2020; Varas-Diaz et al., 2020; Verschueren et al., 2020) (Table 2).

### Meta-analyses and quantitative synthesis

#### Markers of mental fatigue

Both subjective and objective markers were used to evaluate whether mental fatigue was successfully induced in the participants of the included studies. Subjective mental fatigue was evaluated in 9 studies (Boolani et al., 2020; Fletcher & Osler, 2021; Gebel et al., 2022; Morris & Christie, 2020; Nikooharf Salehi et al., 2022; Qu et al., 2020; Tassignon et al., 2020; Varas-Diaz et al., 2020; Verschueren et al., 2020). However, only six studies reported adequate descriptive data and were included in meta-analyses (Boolani et al., 2020; Morris & Christie, 2020; Tassignon et al., 2020; Verschueren et al., 2020). Because, two out of these six studies involve both young and older adults, 8 data sets (four each in young and older adults) were pooled in meta-analyses. Additionally, because the subjective feeling of mental fatigue was measured on scales with variable minimum and maximum scores in the studies, a standardized mean difference in the subjective feeling of mental fatigue between the experimental (mental fatigue) and control conditions was estimated for these studies. The result of the meta-analyses revealed a significantly higher subjective feeling of mental fatigue after the experiment compared to the control condition in both young and older adults (young adults: SMD 1.08, 95% CI 0.36 to 1.79, p-value = 0.003; older adults: SMD 1.79, 95% CI -0.01 to 3.59, p-value = 0.05) (Fig. 4a&b). Considerable heterogeneity (I^2^ = 93 %, p < 0.00001) was observed between the included studies in older adults, indicating a significant variation in subjective mental fatigue outcome between the studies.

**Fig 4.**
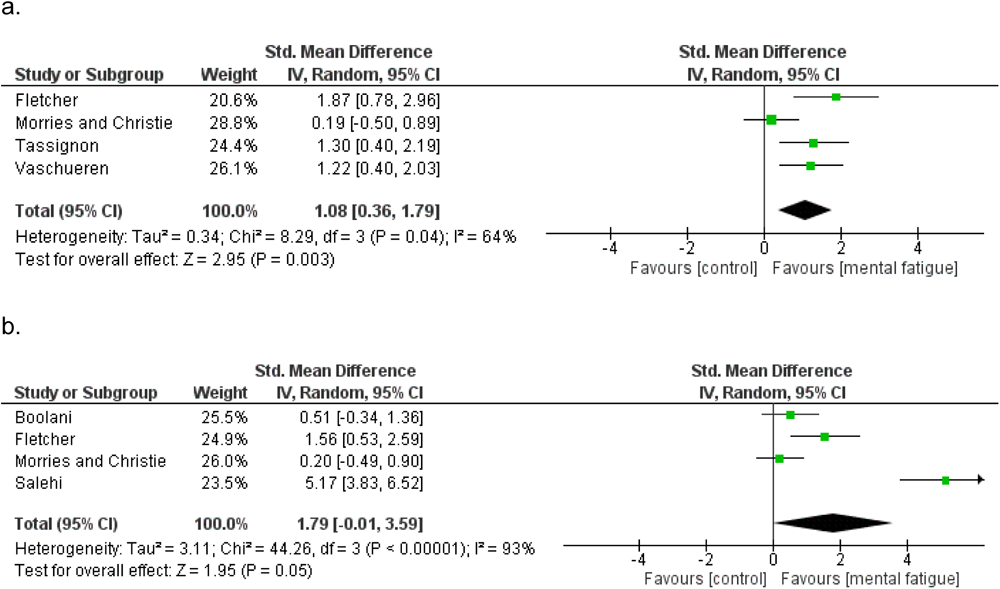
a & b. Meta-analysis results showing the difference in subjective mental fatigue between the experimental and control conditions in a) young and b) older adults.

Among the remaining three studies that evaluated subjective mental fatigue but were not included in the meta-analysis due to a lack of reporting of descriptive data, one study found a significantly higher feeling of mental fatigue after the experimental compared to the control task in older adults (Varas-Diaz et al., 2020). The remaining two studies found a significant increase in subjective mental fatigue post-experimental task compared to pre-experimental task in young adults (Gebel et al., 2022; Qu et al., 2020). The rest of the studies (3 studies) included in this review did not assess subjective mental fatigue (Deschamps et al., 2013; Hachard et al., 2020; Lew & Qu, 2014).

Objectively, mental fatigue due to prolonged cognitive tasks is often defined in terms of the deterioration in the indices of cognitive task performance (reaction time and accuracy) between the beginning (first block) and the end (last block) of the task. Six studies reported change in the indices of cognitive performance between the first and the last blocks of the fatigue-inducing cognitive tasks in young adults and five of them were included in the meta-analysis (Deschamps et al., 2013; Lew & Qu, 2014; Morris & Christie, 2020; Tassignon et al., 2020; Verschueren et al., 2020). The remaining study did not report descriptive data and therefore was not included in the meta-analysis (Hachard et al., 2020).

Four studies reported the change in reaction time (in milliseconds-ms) between the first and last blocks of the cognitive tasks, and these were pooled in a meta-analysis. The result of the meta-analysis revealed a significant increase in reaction time in the last compared to the first block of the cognitive task (MD 19.75 ms, 95%CI 2.71 to 36.79, p-value 0.02) (Fig. 5a). Two studies reported the change in the percentage of accurate responses between the first and last blocks of the cognitive task, while another study reported the error rate difference between the first and the last blocks of the task. Meta-analysis of the accuracy data pooled from the three studies revealed no significant difference in accuracy between the first and final blocks of the cognitive task (SMD -2.15, 95% CI -4.66 to 0.35, p-value = 0.09) (Fig. 5b). Substantial heterogeneity was observed between studies that assessed accuracy (I^2^ = 95%, P < 0.00001), possibly because one of the studies used a cognitive task and accuracy measure that differed from that of the other two studies.

**Fig 5.**
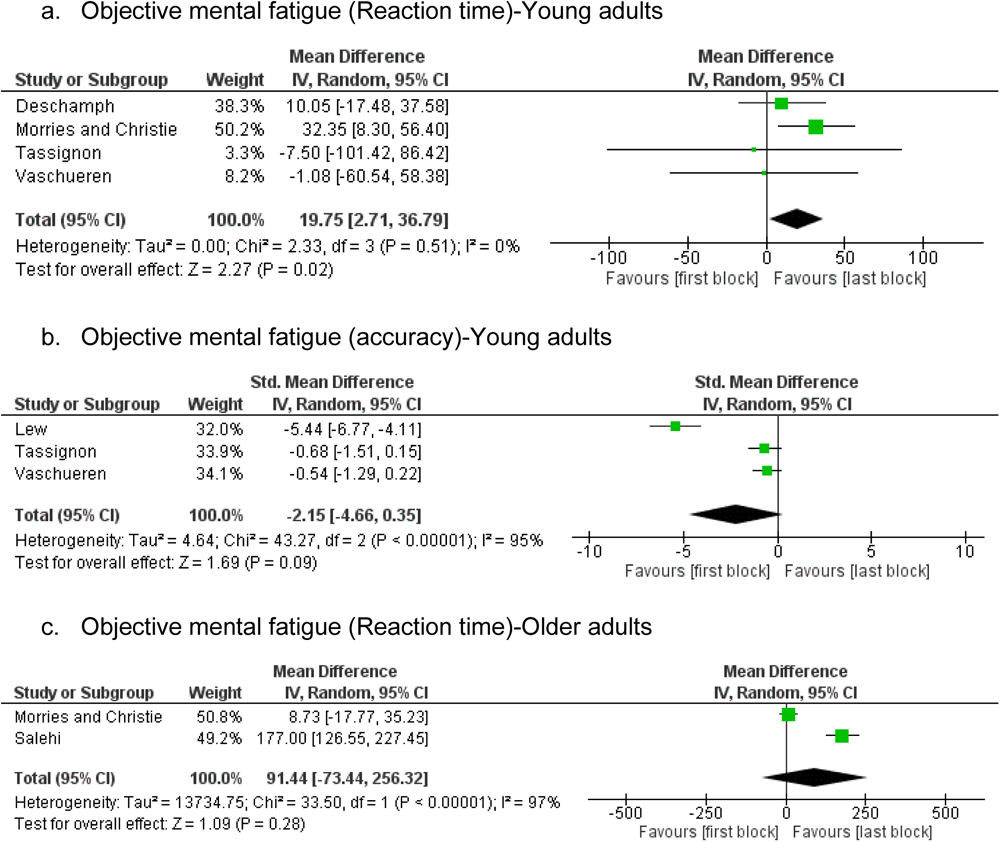
a, b & c. Meta-analysis results showing the difference in cognitive performance between the first and last blocks of the cognitive task in young and older adults (objective markers of mental fatigue).

In older adults, two studies reported the change in reaction time (in milliseconds-ms) between the first and last blocks of the cognitive tasks and were pooled in a meta-analysis (Morris & Christie, 2020; Nikooharf Salehi et al., 2022). Result of the meta-analysis revealed that there was no significant increase in reaction time in the last compared to the first block of the cognitive task (MD = 91.44, 95% CI -73.44 to 256.32, p-value = 0.22) (Fig. 5c). Considerable heterogeneity (I^2^ = 97 %, p < 0.00001) was observed between these studies, indicating a significant variation in objective mental fatigue outcome between the studies.

### Effect of mental fatigue on Balance control in young adults

#### Steady-state balance

Two high quality studies and one low quality study investigated the effect of mental fatigue on steady-state balance involving double-leg stance, in young adults (Deschamps et al., 2013; Fletcher & Osler, 2021; Hachard et al., 2020). However, only two studies reported adequate descriptive data and were included in the meta-analysis (Deschamps et al., 2013; Fletcher & Osler, 2021). Centre of pressure (COP) sway area and total sway path length were used as the outcome measures in the two studies. Result of the meta-analysis revealed no significant difference in COP sway between the mental fatigue and control conditions (SMD = 0.34, 95% CI -0.29 to 0.96, p-value = 0.29) (Fig. 6). In one of the two studies included in the meta-analysis (Fletcher & Osler, 2021), balance was also assessed under dual task condition, and effect size was calculated for the dual task condition too (MD = -0.61, 95% CI -6.86 to 5.64, p-value = 0.85). In addition to quiet standing, one of the three studies also investigated the effect of mental fatigue on Timed-up-and-go (TUG) test time under single and dual task conditions in young adults (Fletcher & Osler, 2021). Effect sizes calculated from the data in this study did not reveal any significant change in TUG time due to mental fatigue (Single task: MD = 0.14, 95% CI -0.24 to 0.52, p-value = 0.47; Dual task: MD = -0.05, 95% CI -0.61 to 0.51, p-value = 0.86).

**Fig 6.**
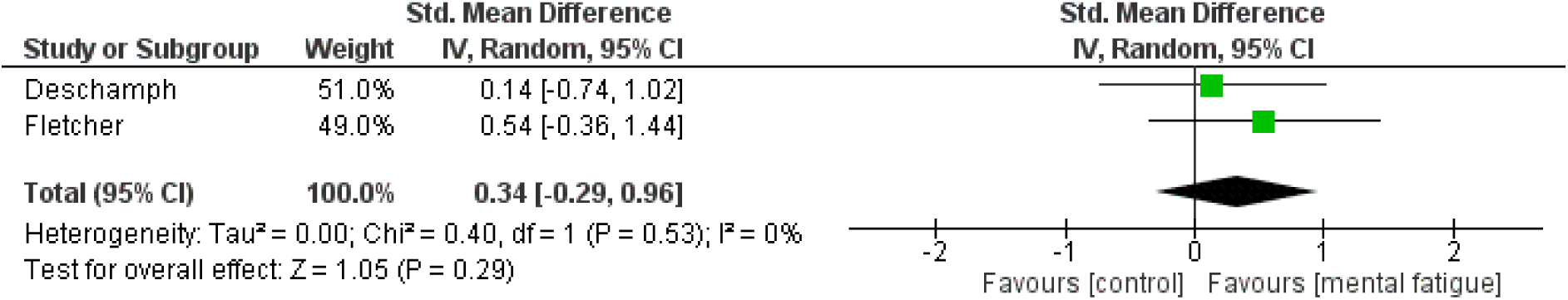
Meta-analysis results showing the effect of mental fatigue on steady-state balance in young adults.

### Proactive (anticipatory) balance

Three high quality and one low quality studies investigated the effect of mental fatigue on measures of proactive balance in young adults (Gebel et al., 2022; Hachard et al., 2020; Tassignon et al., 2020; Verschueren et al., 2020). Two studies used Y-balance test (YBT) and its neurocognitive variant (neurocognitive YBT) (Tassignon et al., 2020; Verschueren et al., 2020), while the remaining two studies measured COP sway while the participants were controlling balance on a wobble board (Gebel et al., 2022; Hachard et al., 2020). The data from the studies that used YBT were pooled in a meta-analysis, while effect sizes were calculated for one of the studies that reported COP sway (Gebel et al., 2022), as the other study did not report adequate descriptive data (Hachard et al., 2020). There was no significant effect of mental fatigue on any of the YBT measures (Fig. 7a-c). Similarly, there was no significant effect of mental fatigue on COP sway while balancing on the wobble board (COP displacement: MD = -16.86, 95% CI -136.4 to 102.5, p-value = 0.78; COP velocity: MD = -0.07, 95% CI -0.98 to 0.84, p-value = 0.84). However, the participants accuracy on the neurocognitive YBT was lower after the mental fatigue compared to the control condition (MD = -5.47, 95% CI -8.68 to -2.27, p-value = 0.0008) (Fig. 7e).

**Fig 7.**
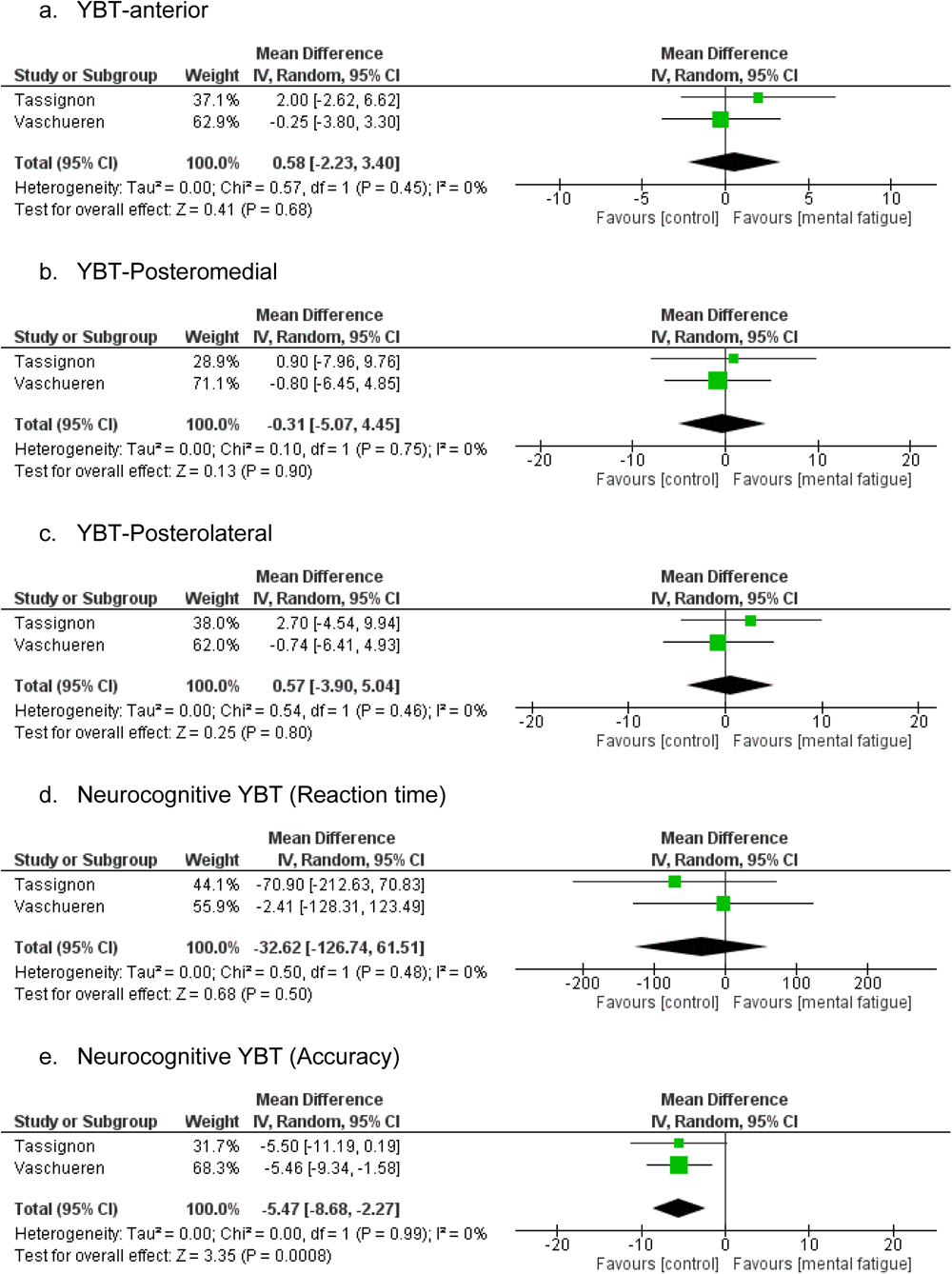
a, b, c, d & e. Meta-analyses results showing the effect of mental fatigue on Proactive (anticipatory) balance control in young adults.

### Reactive balance

Three high quality studies investigated the effect of mental fatigue on reactive balance in young adults (Lew & Qu, 2014; Morris & Christie, 2020; Qu et al., 2020). In two studies, the reactive balance task involves the recovery response to slip and trip during walking (Lew & Qu, 2014; Qu et al., 2020), while the remaining one study involved balance response to unexpected perturbation while participants were standing on a force platform (Morris & Christie, 2020). Meta-analyses of these studies revealed a significant negative effect of mental fatigue on the measures of centre of mass (COM)/ centre of pressure (COP) displacement and velocity used in the studies (displacement: SMD = 0.62, 95% CI 0.27 to 0.97, p-value = 0.0005; velocity: SMD = 0.42, 95% CI 0.00 to 0.85, p-value = 0.05) (Fig. 8a & b).

**Fig 8.**
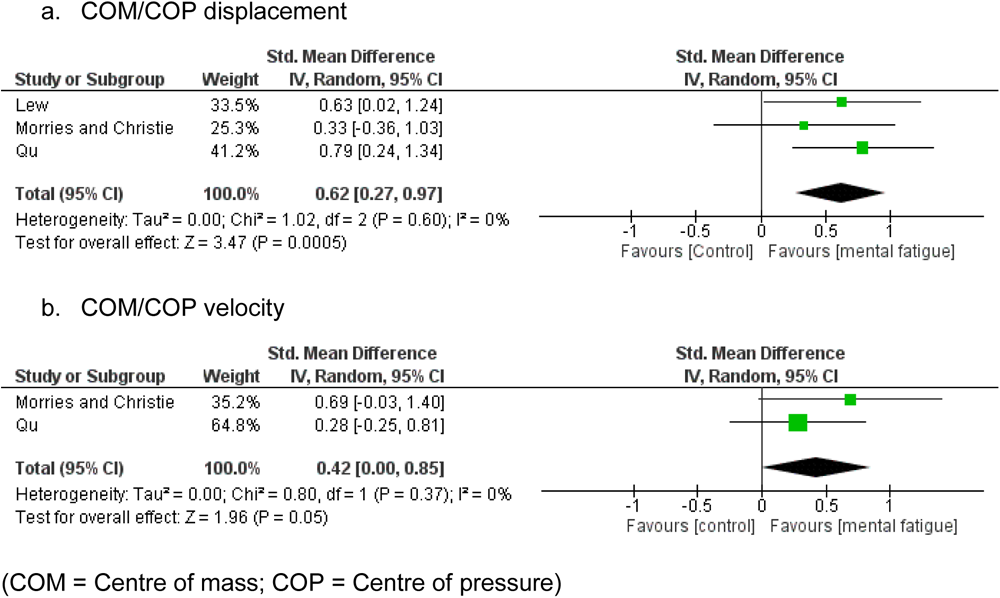
a & b. Meta-analyses results showing the effect of mental fatigue on reactive balance control in young adults.

### Effect of mental fatigue on Balance control in older adults

#### Steady-state balance

The effect of mental fatigue on steady-state balance in older adults was investigated in one high quality study during double-leg stance (Fletcher & Osler, 2021). Balance was assessed under both single and dual tasks conditions by measuring COP sway (Total COP path length) in this study. Effect sizes calculated from this study revealed a significant negative effect of mental fatigue on steady-state balance under single task condition, but not under dual task (Single task: MD = 8.66, 95% CI 1.37 to 15.95, p-value = 0.02; Dual task: MD = 2.82, 95% CI -6.24 to 11.88, p-value = 0.54). Two high quality studies investigated the effect of mental fatigue on Timed-up-and-go (TUG) test time in older adults and were pooled in a meta-analysis (Boolani et al., 2020; Fletcher & Osler, 2021). In one of these studies, the TUG was also conducted under dual task in addition to single task (Fletcher & Osler, 2021). There was no significant effect of mental fatigue on TUG time (MD = 0.57, 95% CI -0.05 to 1.19, p-value = 0.07) (Fig. 9). Effect size calculated from the TUG time during dual tasking does not reveal any significant effect of mental fatigue (MD = 0.58, 95% CI -0.4 to 1.56, p-value = 0.25).

**Fig 9.**
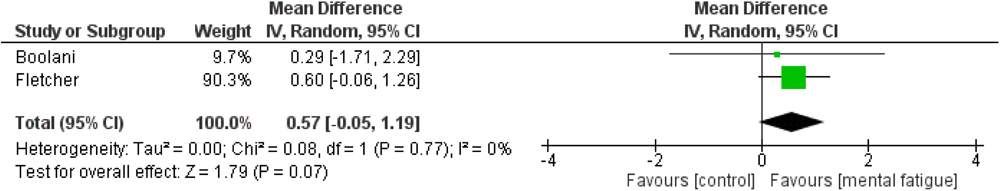
Meta-analysis results showing the effect of mental fatigue on steady-state balance in older adults.

### Proactive (anticipatory) balance

One high quality study investigated the effect of mental fatigue on proactive balance in older adults (Nikooharf Salehi et al., 2022). Participants in this study were asked to maintain balance on the Biodex balance system platform set at medium level of stability (level 6). Mental fatigue led to significant decrease in the different measures of proactive balance used in this study (Overall stability index: MD = 2.74, 95% CI 1.88 to 3.60, p < 0.00001; Anterior/posterior stability index: MD = 2.74, 95% CI 1.90 to 3.58, p < 0.00001; Medial/lateral stability index: MD = 2.74, 95% CI 1.89 to 3.59, p < 0.00001).

### Reactive balance

Two high quality studies investigated the effect of mental fatigue on reactive balance response to unexpected perturbation while standing in older adults (Morris & Christie, 2020; Varas-Diaz et al., 2020). Measures of COM/COP velocity were used as outcomes in these studies. Result of the meta-analysis involving these studies did not reveal significant effect of mental fatigue (SMD = 0.18, 95% CI -0.33 to 0.69, p = 0.49) (Fig. 10).

**Fig 10:**
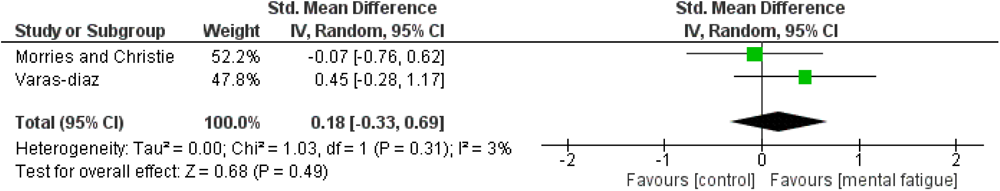
Meta-analysis results showing the effect of mental fatigue on reactive balance control in older adults.

## Discussion

This systematic review and meta-analysis were conducted to synthesize the available evidence relating to how different types of balance tasks are affected by mental fatigue in young and older adults. Prior to delving into the impact of mental fatigue on balance performance, it is imperative to examine the discoveries pertaining to the effective induction of mental fatigue among the participants in the included studies. This is because, in the absence of a successful induction of mental fatigue, it is not possible to confidently attribute any alterations in balance performance observed in the participants to mental fatigue. Successful induction of experimental mental fatigue has been defined using both subjective scales and objective indices related to cognitive performance, such as reaction time and accuracy rate (Chen et al., 2020; Holtzer et al., 2010; Shigihara et al., 2013). The meta-analysis results in this study revealed a significantly higher feeling of mental fatigue after the experimental (mental fatigue) compared to the control condition in both young older adults. Regarding the objective markers of mental fatigue, younger adults exhibit a longer reaction time in the last compared to the first block of the mental fatigue tasks in the included studies, indicating objective evidence of mental fatigue (drop in cognitive performance) in addition to subjective feeling of mental fatigue (Shigihara et al., 2013). Contrarily, there was no significant change in performance between the first and last blocks of the cognitive tasks used to induce mental fatigue in older adults. The meta-analysis concerning objective mental fatigue in the older adults comprises only two studies and relatively smaller number of participants. Consequently, the absence of a significant effect could be attributed to the limited power of the analysis, stemming from the restricted number of studies and participants involved (Haas, 2012). Indeed, the effect direction also indicated a longer reaction time in the last block of the cognitive tasks compared to the first block in this population. This finding suggests that objective mental fatigue, characterized by a decline in cognitive performance, may also be observed if the number of participants in the older age group is increased.

Overall, there is evidence of successfully inducing mental fatigue among the participants in the included studies, particularly in terms of the subjective feeling of mental fatigue, which was observed in both the young and older participants. As a result, mental fatigue might be accountable for the observed changes in the performance of certain balance tasks within this systematic review and meta-analysis.

The results of the meta-analysis (as well as the effect sizes calculated from the remaining studies not included in meta-analysis) on the effect of mental fatigue on steady-state balance control tasks in young adults revealed no significant effect of mental fatigue. Therefore, steady-state balance may not be negatively affected by mental fatigue in this population. The performance of steady-state balance control tasks, such as double-leg stance or walking at a constant velocity, is familiar and well-learned among young adults. These tasks depend on the automatic process of balance control, which is governed by postural reflexes, along with proper body alignment and muscle tone (Jacobs & Horak, 2007; Takakusaki, 2017). The lack of cognitive involvement in the control of these balance tasks in young adults may explain why they are unaffected by mental fatigue (Bernard-Demanze et al., 2009; Boisgontier et al., 2013). On the contrary, in older adults, the effect size calculated from one study examining the impact of mental fatigue during steady-state balance revealed a significant increase in postural sway during double-leg stance due to mental fatigue in this population. However, a meta-analysis encompassing an additional two studies demonstrated that mental fatigue does not have a significant impact on the Time Up and Go (TUG) test (another form of steady-state balance task involving walking) among older adults. The TUG time used as a measure of balance in these studies may not be sensitive enough to detect subtle changes in balance due to mental fatigue (Zampieri et al., 2010). If the participants were equipped with an instrument capable of measuring postural sway, a similar increase in sway might be observed during the TUG test (Zampieri et al., 2010). This assertion is however speculative and should be tested in future studies. Alternatively, the length or difficulty of the cognitive tasks was not great enough to induce significant mental fatigue that can affect balance in the studies that assessed balance using the TUG test. Nonetheless, the observed increase in postural sway during a steady-state balance task (double leg stance) among older adults may be attributed to age-related declines in the structure and function of the various systems involved in balance control, including the musculoskeletal, visual, vestibular, and somatosensory systems, in comparison to younger adults (Horak et al., 1989; Vandervoort, 2002) (Boisgontier et al., 2013; Glasser & Campbell, 1998; Horak et al., 1989; Marsh & Geel, 2000; Rosenhall, 1973; Teasdale et al., 1993; Vandervoort, 2002). Consequently, older adults may require cognitive processing to control steady-state balance tasks that would typically be regulated automatically by brainstem and spinal cord neural circuits (Boisgontier et al., 2013; Marsh & Geel, 2000; Teasdale et al., 1993). This increased reliance on cognitive processes could be the reason why steady-state balance tasks become more susceptible to cognitive manipulations, such as mental fatigue, within this population. However, since balance abilities vary among older adults (Fujita et al., 2015), the susceptibility of steady-state balance tasks to the effects of mental fatigue may also differ within this population, depending on their individual abilities. Older adults displaying signs of frailty may potentially experience a more pronounced increase in postural sway during steady-state balance tasks due to mental fatigue. This intriguing question warrants further investigation in future studies.

Like the steady-state balance, proactive balance control while standing on wobble board or carrying out YBT was also not affected by mental fatigue in young adults. However, result of the meta-analysis revealed significant decrease in accuracy while carrying out the neurocognitive YBT which is also a form of proactive balance task. Unlike the ordinary YBT, where participants had to push reach indicators along three axes (anterior, posteromedial, and posterolateral) as far as possible, while maintaining balance on their dominant leg (Verschueren et al., 2020), the neurocognitive YBT involves reacting as fast as possible with one foot while standing on the contralateral foot to extinguish LED-lights of different colours placed at 80% of the participants’ maximal reach distance in the three axes of the YBT (Verschueren et al., 2019). The colours of the LED-light representing the three axes were informed to the participants beforehand, and they are required to respond and extinguish LED in a particular axis when the colour representing that axis appear in another LED located in front of them (Verschueren et al., 2019). This arrangement in the neurocognitive YBT, whereby a person is required to memorize the different colours corresponding to each axis, recall them, and respond quickly, adds to the cognitive load of this test (Verschueren et al., 2020). Accordingly, although proactive balance control involves goal-directed movements that require planning, which is a form of higher-order cognitive function, young adults might have enough remaining cognitive resources to successfully carry out these tasks under low cognitive load even when they are mentally fatigued. However, when the proactive balance task involves extra cognitive load as seen with the neurocognitive YBT, it became more susceptible to mental fatigue in this population (Tassignon et al., 2020; Verschueren et al., 2020). Possibly, proactive balance control in young adults is more susceptible to the effect of mental fatigue under dual task conditions since the neurocognitive YBT is in a way like dual tasking (increased cognitive load). However, this hypothesis needs to be investigated in future studies. In the older adults, the effect sizes calculated from the single study that investigated the effect of mental fatigue on proactive balance in older adults have revealed a significant negative effect of mental fatigue on all the outcome measures used. Since this study assessed the proactive balance under single task condition, it is plausible to suggest, that unlike in younger adults where proactive balance is only affected by mental fatigue during dual tasking, similar type of balance is affected by mental fatigue even under single task conditions in older adults. This finding can also be explained by the age-related increase in the involvement of attentional resources for the control of balance (Boisgontier et al., 2013; Marsh & Geel, 2000; Salihu et al., 2022; Teasdale et al., 1993). However, as already stated, our finding in older adults was based on a report of a single study which limits the ability to make strong conclusions.

The last type of balance control tasks investigated in this systematic review is the reactive balance control tasks. Result of the meta-analyses revealed a significant effect of mental fatigue on these types of balance tasks in young adults. A significant increase in postural sway was observed in the mental fatigue condition compared to the control condition, indicating an insufficient recovery response and an elevated risk of falling from externally delivered perturbations due to mental fatigue (Lew & Qu, 2014; Qu et al., 2020). Reactive response to balance perturbations involves a continuum of movements ranging from ankle through hip motion (fixed-support strategies), as well as a change in support strategy by taking a step or reaching out to hold on to something with hand to restore balance and prevent fall (Jacobs & Horak, 2007; Shumway-Cook & Woollacott, 2007). The cognitive involvement in balance control increases with increasing novelty or challenge of the balance task (Lajoie et al., 1993). Reactive balance response to unexpected perturbations, such as those that occur due to slips or trips, are quite novel and challenging, and would require active cognitive involvement, even in young adults, especially when they are large enough to require taking a step or holding on to something close by to restore balance (Jacobs & Horak, 2007; Takakusaki, 2017). Several studies suggest the involvement of higher cortical functions, including attention and other executive functions, in ensuring optimal response throughout the various stages involve in reactive balance response to unexpected perturbation (Jacobs & Horak, 2007; Maki & McIlroy, 2007). It was suggested that the cerebral cortex likely influences balance responses to unexpected perturbation both directly, via the corticospinal loops, and indirectly by communicating with the brainstem centres that harbor the synergies for postural responses (Jacobs & Horak, 2007). The deficient reactive balance response observed in young adults due to mental fatigue, as identified in this systematic review and meta-analysis, therefore, aligns with the existing evidence that supports the connection between balance control and higher-order cognitive processing, particularly in situations where the balance task is novel or challenging (Jacobs & Horak, 2007; Lajoie et al., 1993; Maki & McIlroy, 2007). Importantly, since most fall incidents occur due to trips or slips (Ashari et al., 2021; Heijnen & Rietdyk, 2016; Wang et al., 2021), mental fatigue may be an important risk factor increasing the likelihood of fall through its effect on reactive balance control even in young population. During the reactive balance control tasks, we anticipated a stronger negative effect of mental fatigue in older adults compared to young adults. However, contrary to our expectations, the meta-analysis result did not reveal a significant impact of mental fatigue on reactive balance in older adults. It is evident that the number of studies, outcomes, and participants included in the meta-analysis on the effect of mental fatigue on reactive balance in older adults is lower than that of the younger adults. Consequently, the reduced number of studies involving older adults within this review, along with the smaller participant size in these studies, may have diminished the statistical power of our analysis to detect significant results (Haas, 2012). Alternatively, the introduction of unexpected perturbation while standing in the studies involving older adults, in contrast to unexpected slip or trip during walking used in most of the studies involving young adults, may be responsible for the different findings (Lew & Qu, 2014; Qu et al., 2020).

## Conclusion

Investigation on the effect of mentally fatiguing cognitive tasks on balance is an emerging area of research. Most studies have included young participants, and this review determined that mental fatigue impairs proactive balance control under increased cognitive load in this population. Similarly, reactive balance to an unexpected perturbation was affected by mental fatigue. Therefore, it is plausible that mental fatigue may predispose young adults to a loss of balance, falls and possible injury during these types of balance control tasks. In the older population, it was demonstrated that mental fatigue may affect balance during simple steady-state balance tasks, such as double leg stance. However, this finding was based on a report of a single study, which limits the ability to make strong conclusions. It is evident that there is a redistribution of attentional resources as part of the ageing process, therefore, further investigation is required to determine whether mental fatigue adds to the falls risk, which is greater in older people.

### Limitations

This review included a limited number of studies, particularly when considering the older adults, which restricts the capacity to draw robust conclusions due to the small number of participants available for pooling in the meta-analysis. Indeed, some of the findings were derived from effect sizes calculated from a single study, which further constrains the ability to draw strong conclusions.

### Direction for future research

There have been few studies that investigated the effect of mental fatigue on balance, and they have mostly included a small number of participants, the majority of whom were healthy and younger. The people most vulnerable to sustaining a fall related injury are older adults, specifically those with signs of frailty, and people who have suffered injury or disease with a permanent physical and cognitive disability. Therefore, future investigations need to consider each or all these factors to be able to determine both the effect of mental fatigue on balance, and to be able to generalise the findings to vulnerable populations. The taxonomic classification of the difficulty, type, and duration of fatiguing cognitive tasks needs expansion for a more straightforward determination of which combination results in the greater decrement in balance. Additionally, it is crucial to specify the type of balance tasks in which the participants were engaged and to determine the specific effect of mental fatigue on each type of balance task. This will allow for the development of testing and assessment protocols which are more sensitive to detecting changes in balance due to mental fatigue across the spectrum of age and severity of disability.

### Clinical implications

Mental fatigue is highly prevalent in various occupational settings, particularly those that involve prolonged periods of cognitive tasks. Hence, the observed impact of mental fatigue on balance implies its potential contribution to balance loss, falls, and related injuries in the workplace. This crucial factor should be taken into consideration and addressed to enhance safety and reduce fall incidents in occupational settings.

Moreover, sporting activities often encompass a range of challenging and novel balance tasks that necessitate both proactive and reactive balance responses. Young adults form the majority of the age group engaged in both recreational and elite sports. Previous studies have indicated that mental fatigue is common among elite athletes due to factors such as media and sponsorship engagements, repetitive training drills, over-analysis by coaches, constant thoughts about their respective sports, and problem-solving requirements, among others. Consequently, the observed effect of mental fatigue on proactive and reactive balance in young adults implies that it may diminish athletes’ ability to effectively control their balance during sports activities, leading to falls and injuries. Hence, it is imperative for sports medicine clinicians to recognize this significant factor and address it appropriately to reduce the risk of injurious falls in sporting events.

Furthermore, the impact of mental fatigue on balance in older adults suggests that it should be duly considered and addressed when addressing the multitude of factors contributing to a loss of balance and falls within this population. Finally, many chronic conditions, including multiple sclerosis, Parkinson’s disease, stroke, cancer, and others, are associated with substantial mental fatigue, which could potentially contribute to a loss of balance and falls in patients with these conditions. It is therefore important for managing clinicians to assess and effectively address mental fatigue when devising fall prevention strategies for these patient groups.

## Funding

This research did not receive any specific grant from funding agencies in the public, commercial, or not-for-profit sectors.

### Availability of data and materials

All data generated or analyzed during this study is available and can be provided if required.

### Consent to Participate

Not applicable.

### Consent for Publication

Not applicable.

## Authors contribution

Abubakar Tijjani Salihu: Conceptualization, Methodology, Formal Analysis, Data curation, writing-original draft preparation.

Shapour Jaberzadeh: Conceptualization, Methodology, writing-Review, and editing.

Keith D. Hill: Conceptualization, Methodology, writing-Review, and editing.

## Data Availability

All data produced in the present study are available upon reasonable request to the authors

---

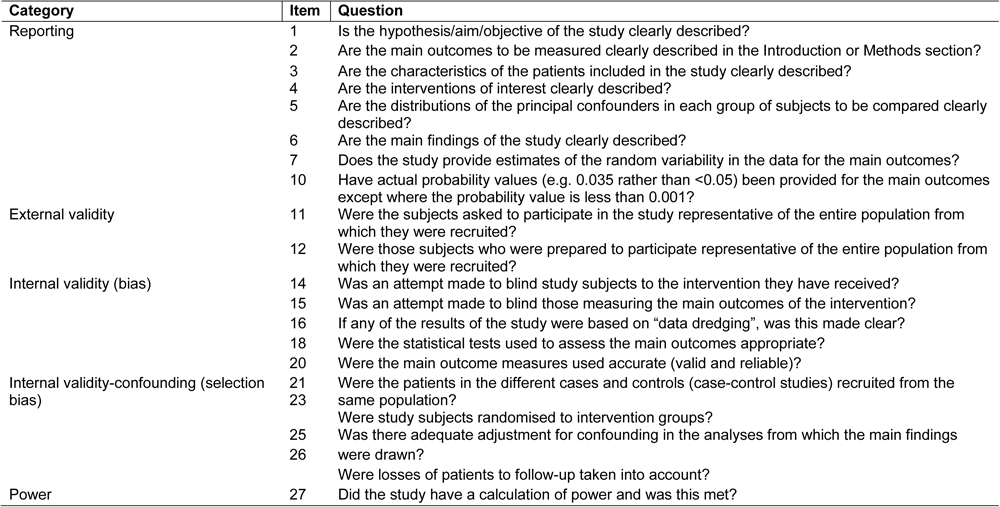
Supplementary material– Modified quality assessment tool as used by Alibazi et al., 2021 & Siddique et al., 2022.

